# Healthcare-related sociocultural factors, racial disparities, and kidney transplant outcomes in the Kidney Transplant Fast Track program

**DOI:** 10.64898/2026.01.16.26344302

**Authors:** Miriam Vélez-Bermúdez, Yuridia Leyva, Chethan Puttarajappa, Arjun Kalaria, Yiliang Zhu, Yue-Harn Ng, Mark L. Unruh, L. Ebony Boulware, Amit Tevar, Mary Amanda Dew, Larissa Myaskovsky

**Author notes:** Corresponding author: Miriam Vélez-Bermúdez, PhD MPH, Center for Healthcare Equity in Kidney Disease, The University of New Mexico Health Sciences Center 901 University Blvd SE STE 150, Albuquerque, NM, 87106.

## Abstract

Background

In the United States, streamlining the kidney transplantation (KT) evaluation process may reduce disparities and barriers to KT access. Prior work showed that the Kidney Transplant Fast Track (KTFT) program shortened this process and reduced racial disparities in waitlisting and overall KT. However, within a setting where evaluation-related structural barriers have been addressed, a comprehensive longitudinal evaluation incorporating sociocultural factors (e.g., medical mistrust, healthcare-related discrimination/racism) alongside race/ethnicity as prespecified predictors across multiple KT milestones, including KT type (living [LDKT] and deceased donor KT [DDKT]), has not been performed.

**Methods:** In this secondary analysis, data came from the KTFT study, a prospective KT candidate cohort. Participants were recruited before KT evaluation start (05/2015-06/2018), coinciding with baseline measure collection, then followed via medical record through 08/2022. We used hierarchically-adjusted Fine-Gray proportional hazards models in this exploratory analysis.

**Results:** Among 1108 KT candidates (243 Black, 783 White, 82 Other), medical mistrust was associated with lower cumulative incidence of waitlisting, but no other sociocultural factors were associated with outcomes. Racial and ethnic differences emerged for KT type: Black participants had a greater cumulative incidence of DDKT, and participants categorized as Other race/ethnicity had a lower cumulative incidence of LDKT, relative to White participants.

Conclusions

Although KTFT reduced racial/ethnic disparities in waitlisting and overall KT receipt, we identified racial/ethnic differences in LDKT and DDKT. Medical mistrust was a significant barrier to waitlisting. Findings suggest that even when the KT evaluation process is streamlined, sociocultural factors and race/ethnicity may influence KT outcomes.

## Introduction

Kidney transplantation (KT) is the optimal therapy for kidney failure.^1–3^ Yet, many patients, particularly those from racially and ethnically minoritized populations in the United States, face persistent barriers to KT access.^3–10^ Before being considered eligible for KT, patients must complete a potentially burdensome, multi-step KT evaluation process spanning several clinical specialties and settings.^11–17^ Typically, the patient is responsible for managing these various appointments and tests across different clinics,^15,16^ and evidence suggests that Black patients take longer than White patients to complete the KT evaluation process,^7^ contributing to broader racial disparities in KT access and receipt.^3^ This disparity may reflect structural barriers across the KT continuum, including upstream delays in referral,^7,11,13–15^ and may be compounded by interpersonal and systemic challenges. Challenges include experiences of discrimination and perceived racism in healthcare, as well as medical mistrust, which Black patients report more frequently than White patients in KT contexts.^7,8,18^

To reduce racial and ethnic disparities in the KT evaluation process, Myaskovsky and colleagues implemented the Kidney Transplant Fast Track (KTFT) program within a large, single-center KT clinic.^19,20^ KTFT is a concierge-based approach that streamlines the KT evaluation process by scheduling all KT patients’ appointments and coordinating receipt of all pre-KT testing.^19,20^ Compared to historical controls, KTFT receipt was associated with greater cumulative incidence of waitlisting and KT after adjusting for sociodemographic and medical characteristics. Moreover, the racial disparity in waitlisting and overall KT receipt (i.e., KT receipt from living and deceased donors) between Black and White patients was no longer significant within the KTFT cohort.^19^

These improvements are especially notable in the context of broader national policy reforms. For example, the 2014 implementation of the Kidney Allocation System (KAS)^21^ allowed patients to begin accruing time on the KT waitlist starting from dialysis initiation, regardless of when they were officially added to the waitlist.^21^ Since KAS implementation,^21^ national trends suggest that racial and ethnic disparities in deceased donor KT (DDKT) have narrowed, but disparities in living donor KT (LDKT) persist.^3–5^ Importantly, KAS did not address barriers in KT evaluation and waitlisting—the stages targeted by KTFT.^19^

Despite the demonstrated success of the streamlining aspect of KTFT in improving racial equity in KT waitlisting and receipt in its patient cohort,^19^ important questions remain. First, we do not yet know whether disparities persisted in the *type of KT received*; the prior study did not examine whether each KT type had racial or ethnic differences.^19^ Given the advantages of LDKT over DDKT, including decreased wait times for KT and increased long-term survival,^22–25^ identifying ongoing disparities in KT type is critical.

Second, although healthcare-related sociocultural factors (henceforth: sociocultural factors), including experiences of discrimination or perceived racism in healthcare, medical mistrust, and trust in physicians, have been identified as key correlates of KT-related outcomes,^7,8,18^ they were not assessed in the prior KTFT study.^19^ Despite their intuitive relevance, their influence on the longitudinal progression through key KT milestones has not been empirically tested within the KTFT cohort,^19^ nor more broadly in settings where structural barriers, like a lengthy KT evaluation process, have been addressed. We conceptualize these four sociocultural factors as interpersonal or systemic experiences within healthcare that can shape patient-provider relationships, trust in healthcare providers and systems, and engagement with complex healthcare processes such as KT evaluation. Prior cross-sectional research across chronic disease contexts demonstrates that these sociocultural experiences are associated with poorer communication, and unmet or delayed care.^7,8,13,26–32^ In previous work, Hamoda and colleagues found that these sociocultural factors, and not race, were associated with lower odds of attending the initial KT evaluation appointment.^8^ This finding was significant, given race has historically served as a proxy for these factors.^33,34^ Unlike race, sociocultural factors may be modifiable targets of racial and ethnic disparities. Thus, examining these factors within the KTFT cohort may reveal whether their influence on key KT-related milestones persists despite reductions in racial disparities, and may provide insights for additional strategies that enhance KT access and equity.

In this exploratory secondary data analysis of a prospective observational cohort, we addressed these two gaps. All participants underwent the KTFT program, which addressed a key barrier of KT by streamlining the evaluation process. KTFT provided the context in which we examined how race and ethnicity, and sociocultural factors, influenced multiple KT outcomes.

We assessed racial and ethnic differences in KT type (LDKT and DDKT) within the KTFT cohort. Also, we examined whether sociocultural factors were associated with multiple time-to-event KT outcomes (i.e., time to active waitlisting, KT receipt, LDKT receipt, and DDKT receipt), after adjusting for demographic, medical, and psychosocial factors, donor recruitment and preference, and transplant knowledge and concerns. Although waitlisting and overall KT receipt were included in the primary KTFT paper as time-to-event outcomes, these models were limited to demographic and medical covariates.^19^ As such, it remains unknown whether observed reductions in racial disparities in waitlisting and KT were primarily driven by streamlining the evaluation process. Therefore, we explored whether sociocultural factors were associated with key KT milestones, and whether they explained, attenuated, or modified racial and ethnic differences in these outcomes. The present analysis introduced a hierarchical modeling framework incorporating an expanded set of variables, representing the first comprehensive evaluation of these sociocultural factors as predictors in longitudinal models across multiple KT outcomes, including KT type, within the KTFT cohort.

## Methods

### Study Sample and Procedures

This prospective observational cohort study of KT candidates is from a larger clinical trial, “Increasing Equity in Transplant Evaluation and Living Donor Kidney Transplantation” (ClinicalTrials.gov identifier: NCT02342119).^19,20^ In the original study, patients were recruited from a single center from 05/2015 through 06/2018. Patients were eligible for recruitment if they were 18 years or older, English-speaking, referred for KT, had not previously undergone KT, did not have cognitive or sensory impairments that would prevent them from participating, and had scheduled their initial KT evaluation appointment.

Among 1,472 eligible patients, we obtained verbal consent for 1,288 patients after their initial KT evaluation appointment was scheduled, but before they attended their clinic appointment. It was during this period that we collected baseline data via telephone interviews or paper surveys. Of the 1,288 who gave verbal consent, only 1,118 attended the initial evaluation clinic appointment. During this appointment, we obtained written informed consent from 1,108 of them to review their medical records. Their waitlisting and KT statuses were followed via medical records through 08/2022. See Myaskovsky et al. 2025 for details on patient flow,^19^ and Bornemann et al. 2017 for detailed information on study protocol.^20^

The current study was approved by the institutional review boards at the University of Pittsburgh and the University of New Mexico, and both institutions signed a data use agreement. Study conduct was in accordance with the Declaration of Helsinki^35^ and consistent with the Principles of the Declaration of Istanbul on Organ Trafficking and Transplant Tourism.^36^ We followed “Strengthening the Reporting of Observational Studies in Epidemiology” reporting guidelines (Supplemental Table 1).^37^

## Study Measures

### Outcome Variables

Outcome variables for this study included the cumulative incidence of 1) active KT waitlisting; 2) KT receipt (DDKT, LDKT, or unknown donor); 3) LDKT receipt; and 4) DDKT receipt.

### Independent Variables

All independent variables were collected at baseline (i.e., after patients’ initial KT evaluation appointment was *scheduled*, but before they *attended* the appointment). The primary independent variables were **race and ethnicity**, and **sociocultural factors**. For race and ethnicity, patients were first asked to report if they were Hispanic or Latino, and then asked their race (i.e., American Indian/Native American, Asian, Black/African American, Native Hawaiian/Pacific Islander, White, and/or “Other” race). We combined race and ethnicity into a race/ethnicity variable that yielded three groups: non-Hispanic Black, non-Hispanic White, and “Other race/ethnicity” (the latter group was created due to the small number of participants who did not indicate either “Black” or “White” race; all other races and ethnicities were collapsed into this category). Sociocultural factors were operationalized with the following four variables: 1) *experience of discrimination in healthcare* (i.e., “discrimination”; 7-item scale reporting personal experiences of discrimination with healthcare providers; response range: “Never” to “Always”),^38,39^ *perceived racism in healthcare* (i.e., “racism”; 4-item scale reporting how common racism is in healthcare; response range: “Strongly disagree” to “Strongly agree”),^40^ *medical mistrust of the healthcare system* (i.e., “mistrust”; 7-item scale reporting how hospital systems are untrustworthy, incompetent, and not acting in patients’ best interest; response range: “Strongly disagree” to “Strongly agree”),^40–42^ and *trust in physician* (i.e., 11-item scale reporting how much respondents trust their physicians; response range: “Totally disagree” to “Totally agree”).^43^ Each measure used a 5-point Likert scale. Items were averaged to create scale scores from 1 to 5, with higher scores reflecting greater discrimination, racism, mistrust, or trust in physician.

We collected self-reported demographic characteristics, transplant knowledge and concerns, psychosocial factors, donation preference and recruitment, and sociocultural factors with the baseline survey. We abstracted baseline medical factors via electronic medical review (see Supplemental Table 2 for a full description of variables).

## Statistical Analysis

We examined our data to identify missingness and ensure analytic assumptions were met. We adjusted independent variables with issues related to normality, skewness, or outliers through either log or squared transformation, or converting from a continuous variable to categorical (e.g., we dichotomized the experiences of discrimination measure into “Ever” and “Never” because its distribution was skewed).^39^ Missing cases for each variable are reported under Table 1, with no observed patterns of missingness by racial/ethnic group. For categorical variables, we calculated frequencies and percentages. For continuous variables, we reported medians and first and third quartiles, which are resistant to potential outliers and closely approximate means for normal distributions. We compared participants by the three race/ethnicity categories on all survey items using Pearson’s Chi-squared Tests or one-way Analysis of Variance (ANOVA) with Sidak correction by race/ethnicity (Table 1).

**Table 1.**
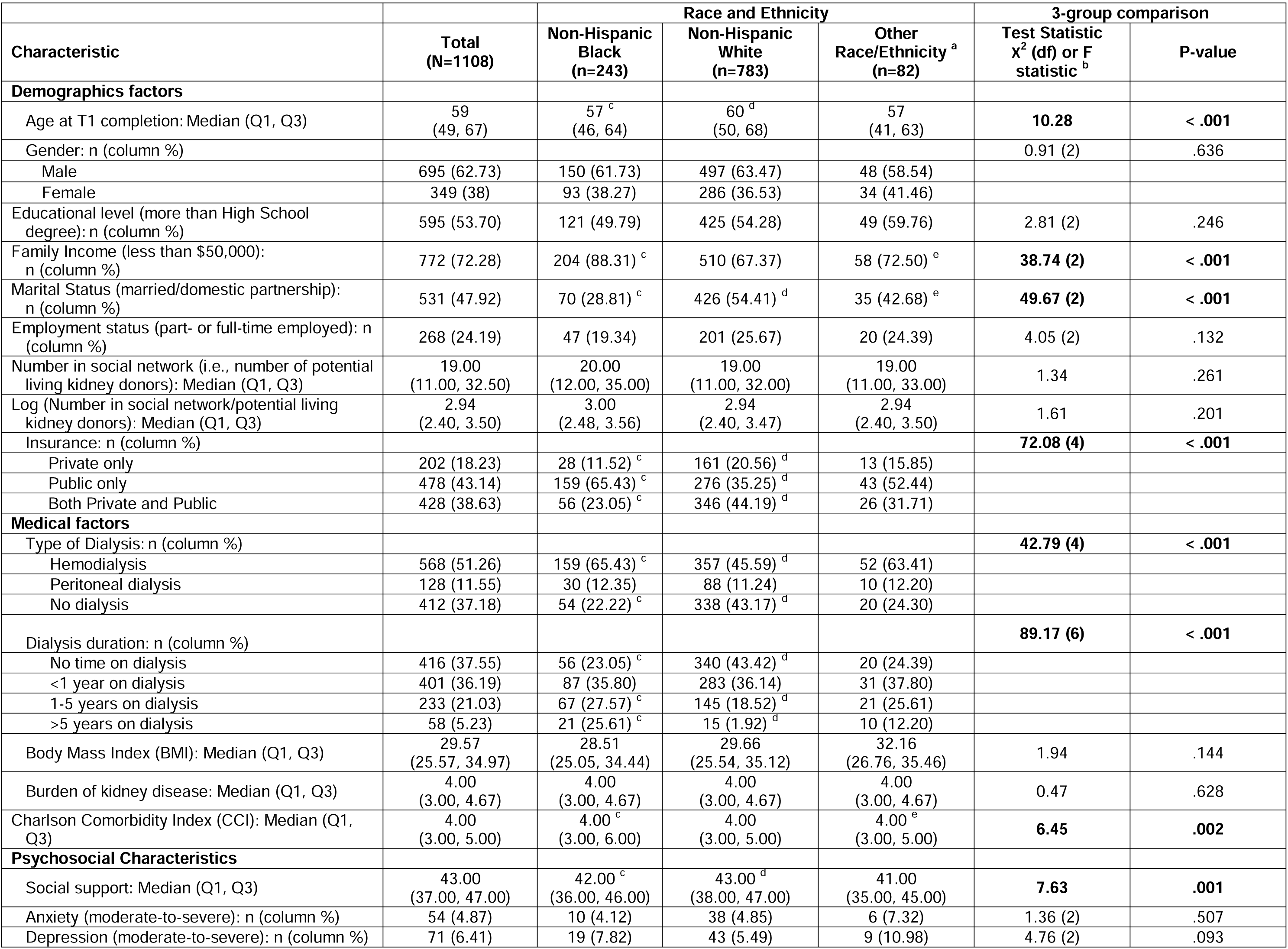
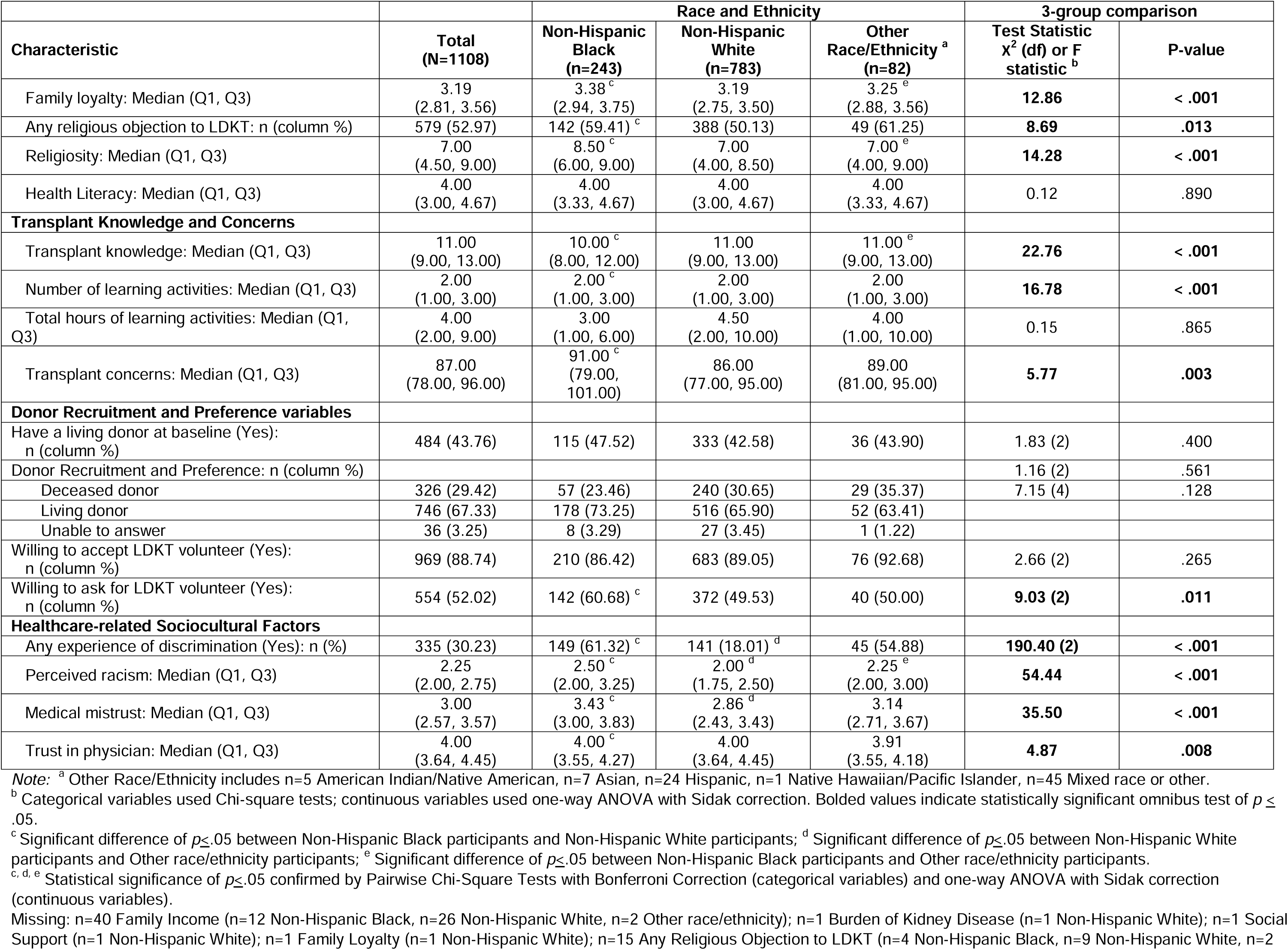
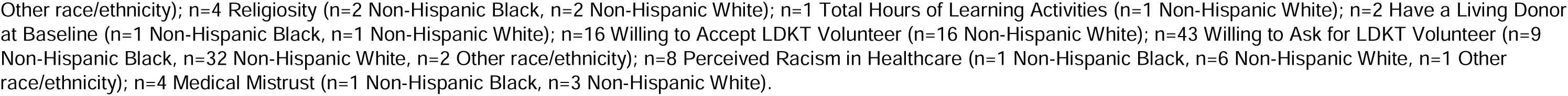
Characteristics of participants at baseline: Total sample, and by race and ethnicity.

### Fine-Gray Proportional Hazards Model

To evaluate race/ethnicity and sociocultural factors as predictors of KT outcomes, we performed Fine-Gray competing risk survival analyses with cumulative incidence function on KT waitlisting, and among those waitlisted, overall KT, LDKT, and DDKT. To determine associations with time to active KT waitlisting, time was measured from evaluation initiation to active waitlisting, regardless of interruptions or restarts, as long as waitlisting occurred before study completion. Censored events included rejection from the waitlist, incomplete evaluation, evaluation closure by patient or clinic, or ongoing evaluations at study completion. Death was categorized as a competing event. To determine associations with time to overall KT, LDKT, or DDKT receipt, time was measured from active waitlisting to KT. Censored events included removal from waitlist or remaining on the waitlist at study completion. Death was a competing event for all models. Because receipt of one KT type precludes receipt of another, DDKT receipt was considered a competing event for the LDKT model, and vice-versa. Given the presence of competing risks, Fine-Gray proportional hazards analysis was preferred over a standard Cox model, as the latter would censor events like death,^44,45^ which is a distinct terminal consequence that prevents active waitlisting or transplantation.

We employed a hierarchical analytic approach^46^ to evaluate associations between race/ethnicity, sociocultural factors, and KT-related outcomes in this exploratory analysis. We first modeled the unadjusted association of race/ethnicity with each outcome to establish baseline differences (Block 1). We then added sociocultural factors to examine their associations with outcomes and evaluate how their inclusion affected race/ethnicity estimates (Block 2). Finally, we added an expanded set of covariates, including demographic, medical, and psychosocial factors; donor recruitment and preference variables; and transplant knowledge or concerns, to evaluate whether race/ethnicity and/or sociocultural factors were associated with outcomes following further adjustment (Block 3). Although our prior publication reported on adjusted time-to-event models with race/ethnicity predicting active waitlisting and overall KT receipt, it only adjusted for demographic and medical characteristics.^19^ The current hierarchical modeling approach allowed us to assess how associations between key predictors (i.e., race/ethnicity, sociocultural factors) and KT outcomes changed across sequentially-adjusted models.

### Model Selection

All variables were considered for inclusion in the fully-adjusted Fine-Gray models across the four outcomes. To determine which covariates to include, we applied the least absolute shrinkage and selection operator (LASSO) method on demographic, medical, and psychosocial factors, transplant knowledge or concerns, and donor preference and recruitment variables.

LASSO applies regularization and shrinkage techniques to identify predictors for model inclusion, reduce overfitting, and address multicollinearity by retaining relevant variables and discarding redundant ones.^47,48^ This LASSO method accounted for competing risks data using the ‘fastcmprsk’ R package.^49^ Based on *a priori* aims, the following variables were forced into the LASSO procedure: race/ethnicity, discrimination, racism, mistrust, and trust in physicians. For each model determined by LASSO, we confirmed the proportional hazards assumption.^50^

## Results

Our study included 1108 patients (male n[%]=695[63]; non-Hispanic Black n[%]=243[22]; non-Hispanic White n[%]=783[71]; Other race/ethnicity n[%]=82[7]). Median age was 59 (interquartile range: 49-67). Participants significantly differed by race and ethnicity on several variables (Table 1). Frequencies and percentages of key KT-related outcomes, including censored and competing events, can be found in footnotes under Tables 2-5.

**Table 2.** Fine-Gray proportional sub-distribution hazards models for time (in days) from initial KT evaluation to active waitlisting (competing event: death)

|  | SHR | p-value | 95% CI |
| --- | --- | --- | --- |
| **Block 1 (unadjusted race/ethnicity model <sup>a)</sup>)** |  |  |  |
| Race (Reference: Non-Hispanic White) |  |  |  |
| Black, non-Hispanic | **0.63** | **< .001** | **0.50 – 0.79** |
| Other race/ethnicity | 0.77 | .121 | 0.56 – 1.07 |
| **Block 2 (race/ethnicity + sociocultural factors <sup>b)</sup>)** |  |  |  |
| Race (Reference: Non-Hispanic White) |  |  |  |
| Black, non-Hispanic | **0.70** | **.005** | **0.55 – 0.90** |
| Other race/ethnicity | 0.80 | .197 | 0.58 – 1.12 |
| Experiences of Discrimination in Healthcare<br>(Reference: Never experienced) | 0.97 | .778 | 0.78 – 1.21 |
| Perceived Racism in Healthcare | 1.04 | .538 | 0.92 – 1.18 |
| Medical Mistrust | **0.79** | **< .001** | **0.69 – 0.90** |
| Trust in Physician | 1.15 | .077 | 0.98 – 1.34 |
| **Block 3 (adjusted model with LASSO-selected covariates <sup>c)</sup>)** |  |  |  |
| * Race (Reference: Non-Hispanic White) |  |  |  |
| Black, non-Hispanic | 0.88 | .361 | 0.68 – 1.15 |
| Other race/ethnicity | 0.80 | .229 | 0.55 – 1.15 |
| * Experiences of Discrimination in Healthcare<br>(Reference: Never experienced) | 1.16 | .191 | 0.93 – 1.46 |
| * Perceived Racism in Healthcare | 1.05 | .426 | 0.93 – 1.19 |
| * Medical Mistrust | **0.85** | **.014** | **0.74 – 0.97** |
| * Trust in Physician | 1.04 | .670 | 0.88 – 1.22 |
| Age | **0.98** | **< .001** | **0.98 – 0.99** |
| Family income of less than \$50k per year<br>(Reference: \$50k or more per year) | 0.85 | .170 | 0.63 – 1.07 |
| Marital status (Single, widowed, or divorced)<br>(Reference: Married or partnered) | **0.68** | **< .001** | **0.55 – 0.84** |
| Employment status (Unemployed)<br>(Reference: Employed full-/part-time) | **0.71** | **.002** | **0.57 – 0.88** |
| Dialysis modality (Reference: Not on dialysis) |  |  |  |
| Hemodialysis | 1.09 | .866 | 0.42 – 2.84 |
| Peritoneal dialysis | 1.06 | .909 | 0.39 – 2.91 |
| Dialysis duration (Reference: No time on dialysis) |  |  |  |
| Less than 1 year on dialysis | 0.69 | .457 | 0.26 – 1.84 |
| 1 – 5 years on dialysis | 0.55 | .251 | 0.21 – 1.51 |
| More than 5 years on dialysis | **0.32** | **.044** | **0.11 – 0.97** |
| CCI | **0.84** | **< .001** | **0.79 – 0.90** |
| Burden of Kidney Disease | **0.91** | **.030** | **0.83 – 0.99** |
| Transplant Knowledge | **1.05** | **.013** | **1.01 – 1.08** |
| Number of Learning Activities | 1.05 | .318 | 0.96 – 1.14 |
| Moderate-to-severe Depression<br>(Reference: No depression) | **0.56** | **.016** | **0.34 – 0.89** |
| Social Support | 1.01 | .090 | 0.99 – 1.03 |

*Note:* Bolded values indicate statistical significance of $p < .05$ . \* Denotes variables that were forced into the LASSO procedure in Block 3 due to *a priori* aims. The remaining variables in Block 3 were selected for inclusion by LASSO. The total study sample was included in this analysis ( $N=1108$ ). The number of events in this analysis was as follows: $n=525$ (47%) participants were actively waitlisted, $n=411$ (37%) were censored, and $n=172$ (16%) experienced a competing event (i.e., death).
<sup>a</sup> See Figure 1A.
<sup>b</sup> See Figure 1B.
<sup>c</sup> See Figure 1C.

### Time to Active Waitlisting

In Block 1, Black participants (sub-distribution hazard ratio [SHR]=0.63; 95% CI: 0.50-0.79; *p*<.001), but not Other race/ethnicity participants (SHR=0.77; 95% CI: 0.56-1.07; *p*=.121), had a significantly lower cumulative incidence of waitlisting compared with White participants. These findings establish baseline racial and ethnic differences within the KTFT cohort prior to further model adjustment. In Block 2, after introducing sociocultural factors, the same statistical pattern was observed (Black participants: SHR=0.70, 95% CI: 0.55-0.90, *p*=.005; Other race/ethnicity participants: SHR=0.80, 95% CI: 0.58-1.12, *p*=.197). In addition, medical mistrust was significantly associated with reduced cumulative incidence of waitlisting (SHR=0.79; 95% CI: 0.69-0.90; *p*<.001). In Block 3, there were no statistically significant racial and ethnic differences regarding time to active waitlisting (Black participants: SHR=0.88, 95% CI: 0.68-1.15, *p*=.361; Other race/ethnicity participants: SHR=0.80, 95% CI: 0.55-1.15, *p*=.229).

However, greater medical mistrust maintained its association with lower cumulative incidence of waitlisting (SHR=0.85; 95% CI: 0.74-0.97; *p*=.014; see Table 2 and Figure 1A-C).

**Figure 1.**
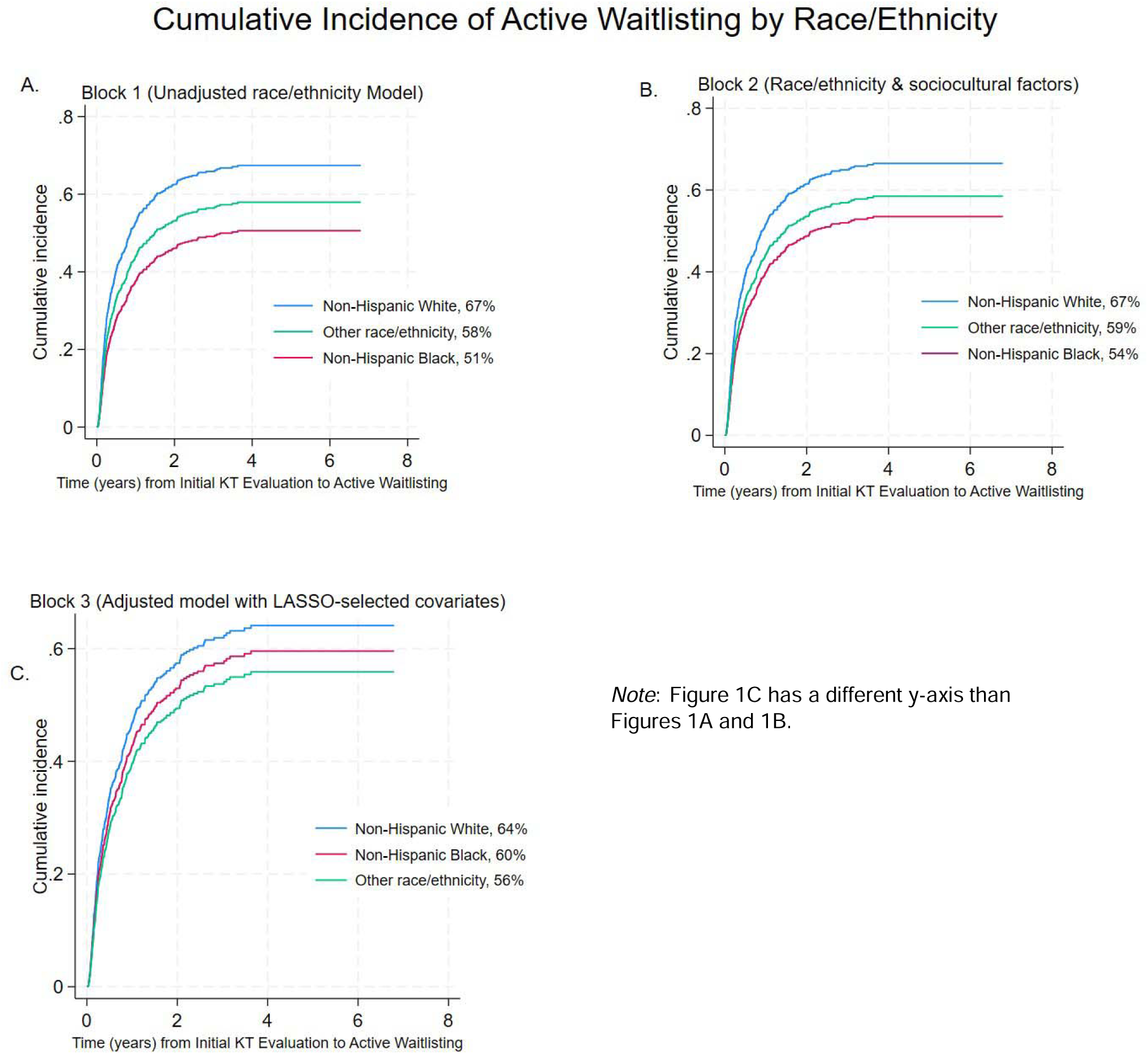
Cumulative incidence of active waitlisting, by race/ethnicity, estimated using Fine-Gray proportional hazard modeling (competing events: Death): Panel A. Block 1 (Unadjusted Model); Panel B. Block 2 (Adjusted for Sociocultural Factors); Panel C. Block 3 (Fully-Adjusted Model).

### Time from Active Waitlisting to KT

In Block 1, there were no statistically significant differences between Black participants (SHR=1.13; 95% CI: 0.86-1.47; *p*=.379) or Other race/ethnicity participants (SHR=1.14; 95% CI: 0.71-1.82; *p*=.592) compared with White participants in the cumulative incidence of KT. This pattern was maintained in Block 2 (Black participants: SHR=1.26, 95% CI: 0.94-1.70, *p*=.128; Other race/ethnicity participants: SHR=1.15, 95% CI: 0.71-1.86, *p*=.563), and Block 3 (Black participants: SHR=1.12, 95% CI: 0.78-1.59, *p*=.541; Other race/ethnicity participants: SHR=1.16, 95% CI: 0.72-1.85, *p*=.548). Furthermore, no sociocultural factors were associated with KT receipt (see Table 3 and Figure 2A-C).

**Figure 2.**
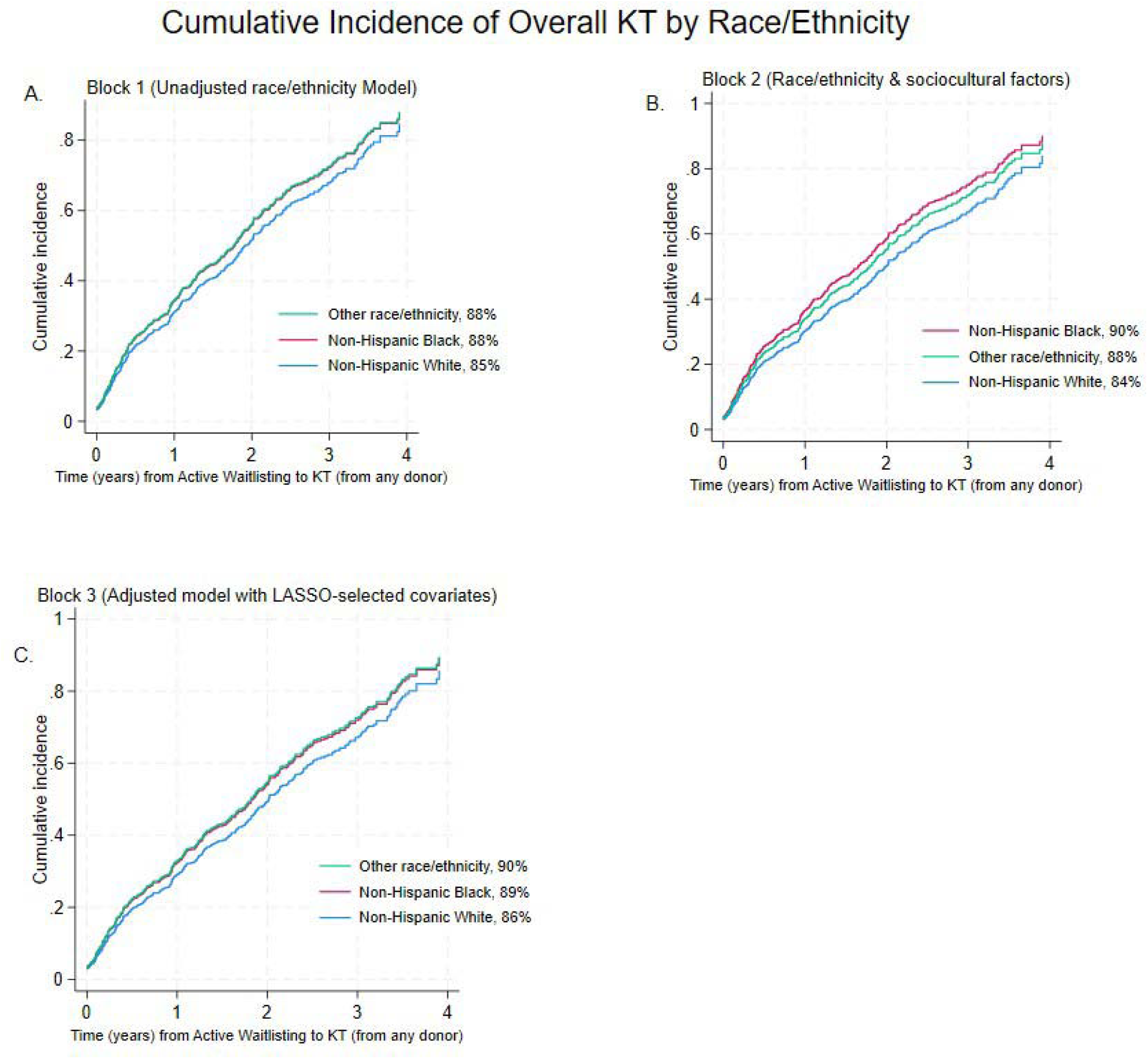
Cumulative incidence of overall KT, by race/ethnicity, estimated using Fine-Gray proportional hazard modeling (competing events: Death): Panel A. Block 1 (Unadjusted Model); Panel B. Block 2 (Adjusted for Sociocultural Factors); Panel C. Block 3 (Fully-Adjusted Model).

**Table 3.** Fine-Gray proportional sub-distribution hazards models for time (in days) from active waitlisting to KT of any type (competing event: death)

|  | SHR | p-value | 95% CI |
| --- | --- | --- | --- |
| **Block 1 (unadjusted race/ethnicity model <sup>a)</sup>)** |  |  |  |
| Race (Reference: Non-Hispanic White) |  |  |  |
| Black, non-Hispanic | 1.13 | .379 | 0.86 – 1.47 |
| Other race/ethnicity | 1.14 | .592 | 0.71 – 1.82 |
| **Block 2 (race/ethnicity + sociocultural factors <sup>b)</sup>)** |  |  |  |
| Race (Reference: Non-Hispanic White) |  |  |  |
| Black, non-Hispanic | 1.26 | .128 | 0.94 – 1.70 |
| Other race/ethnicity | 1.15 | .563 | 0.71 – 1.86 |
| Experiences of Discrimination in Healthcare<br>(Reference: Never experienced) | 0.91 | .478 | 0.69 – 1.19 |
| Perceived Racism in Healthcare | 0.91 | .268 | 0.77 – 1.08 |
| Medical Mistrust | 1.00 | .977 | 0.85 – 1.18 |
| Trust in Physician | 1.07 | .498 | 0.88 – 1.29 |
| **Block 3 (adjusted model with LASSO-selected covariates <sup>c)</sup>)** |  |  |  |
| * Race (Reference: Non-Hispanic White) |  |  |  |
| Black, non-Hispanic | 1.12 | .541 | 0.78 – 1.59 |
| Other race/ethnicity | 1.16 | .548 | 0.72 – 1.85 |
| * Experiences of Discrimination in Healthcare<br>(Reference: Never experienced) | 0.89 | .439 | 0.67 – 1.19 |
| * Perceived Racism in Healthcare | 0.93 | .422 | 0.78 – 1.11 |
| * Medical Mistrust | 1.06 | .491 | 0.90 – 1.25 |
| * Trust in Physician | 1.03 | .774 | 0.84 – 1.26 |
| Age | **0.99** | **.008** | **0.98 – 0.99** |
| Education (High school degree or less)<br>(Reference: More than high school) | 0.82 | .135 | 0.63 – 1.06 |
| Health Insurance<br>(Reference: Public and private insurance) |  |  |  |
| Private insurance only | 1.04 | .767 | 0.78 – 1.39 |
| Public insurance only | 0.88 | .473 | 0.63 – 1.24 |
| Dialysis duration<br>(Reference: No time on dialysis) |  |  |  |
| Less than 1 year on dialysis | 0.79 | .070 | 0.61 – 1.02 |
| 1 – 5 years on dialysis | 1.16 | .421 | 0.81 – 1.67 |
| More than 5 years on dialysis | 2.28 | .088 | 0.88 – 5.86 |
| Health literacy | 1.09 | .266 | 0.94 – 1.27 |
| Any religious objection to LDKT<br>(Reference: No religious objection) | 0.82 | .098 | 0.65 – 1.04 |
| Social Support | 1.02 | .222 | 0.99 – 1.04 |
| Transplant Knowledge | 1.00 | .948 | 0.96 – 1.04 |
| Hours of learning activities | 1.01 | .055 | 0.96 – 1.04 |
| Not having a living donor<br>(Reference: Having a living donor) | **0.77** | **.043** | **0.60 – 0.99** |
| Willing to accept a living donor<br>(Reference: Not willing to accept a living donor) | 0.78 | .219 | 0.52 – 1.16 |

*Note:* Bolded values indicate statistical significance of $p < .05$ . \* Denotes variables that were forced into the LASSO procedure in Block 3 due to *a priori* aims. The remaining variables in Block 3 were selected for inclusion by LASSO. Participants included in this analysis were those actively waitlisted ( $n=525$ ). Among those actively waitlisted, the number of events in this analysis was as follows: $n=330$ (63%) participants were transplanted, $n=142$ (27%) were censored, and $n=53$ (10%) experienced a competing event (i.e., death).
<sup>a</sup> See Figure 2A.
<sup>b</sup> See Figure 2B.
<sup>c</sup> See Figure 2C.

### Time from Active Waitlisting to LDKT

There was a statistically significant lower cumulative incidence of LDKT among Black participants (SHR=0.41; 95% CI: 0.22-0.77; *p*=.005), but not Other race/ethnicity participants (SHR=0.42; 95% CI: 0.16-1.11; *p*=.081), when compared with White participants in Block 1. This pattern persisted in Block 2, although differences were slightly smaller (Black participants: SHR=0.48, 95% CI: 0.26-0.91, *p*=.023; Other race/ethnicity participants: SHR=0.48, 95% CI: 0.18-1.28, *p*=.144). However, in Block 3, the difference between Black and White participants was attenuated for LDKT (SHR=0.47; 95% CI: 0.21-1.03; *p*=.060), but became statistically significant between Other race/ethnicity and White participants (SHR=0.36; 95% CI: 0.14-0.89; *p*=.027). No statistically significant associations between sociocultural factors and LDKT receipt were identified (see Table 4 and Figure 3A-C).

**Figure 3.**
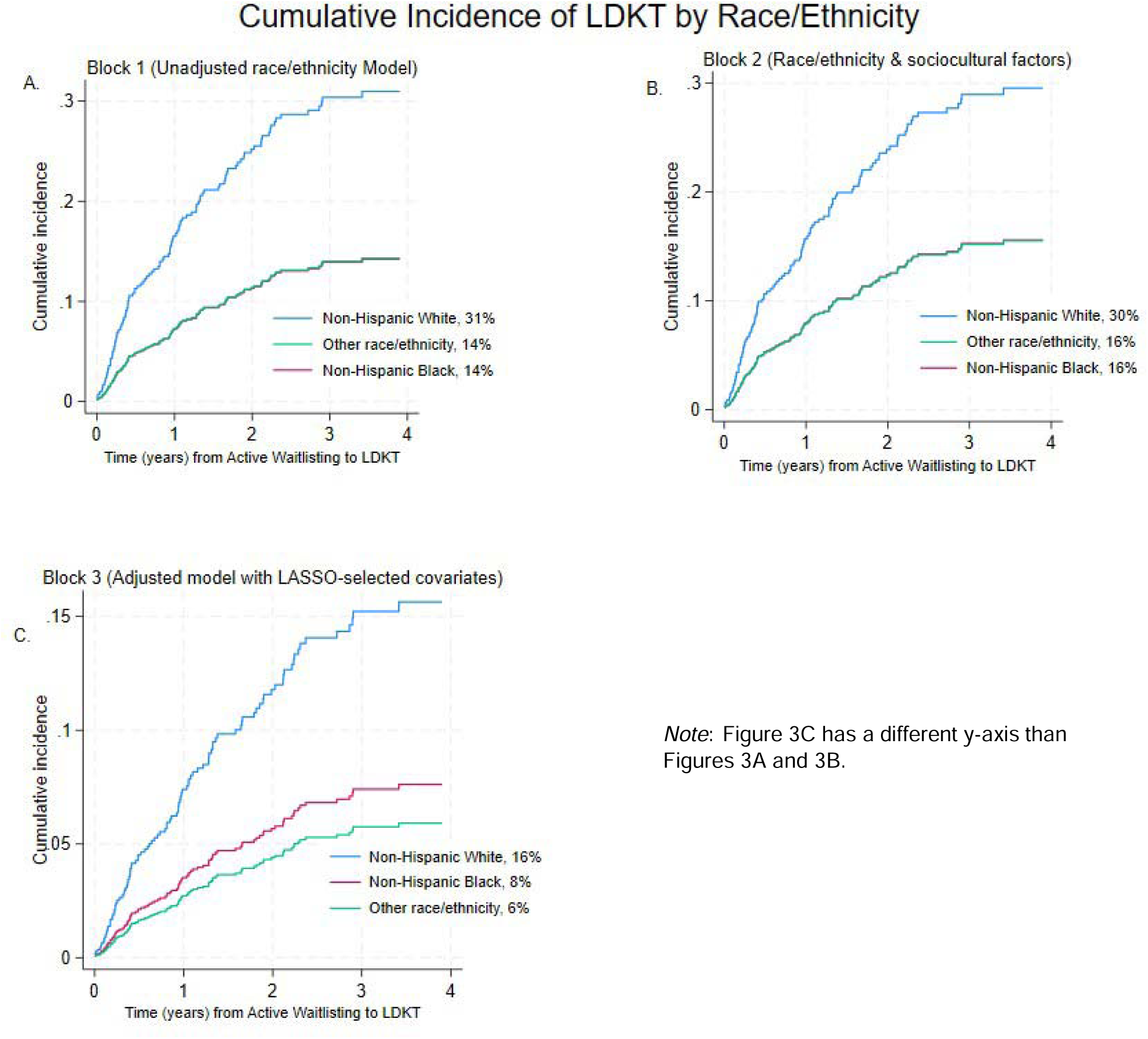
Cumulative incidence of LDKT, by race/ethnicity, estimated using Fine-Gray proportional hazard modeling (competing events: Death, DDKT): Panel A. Block 1 (Unadjusted Model); Panel B. Block 2 (Adjusted for Sociocultural Factors); Panel C. Block 3 (Fully-Adjusted Model).

**Table 4.**
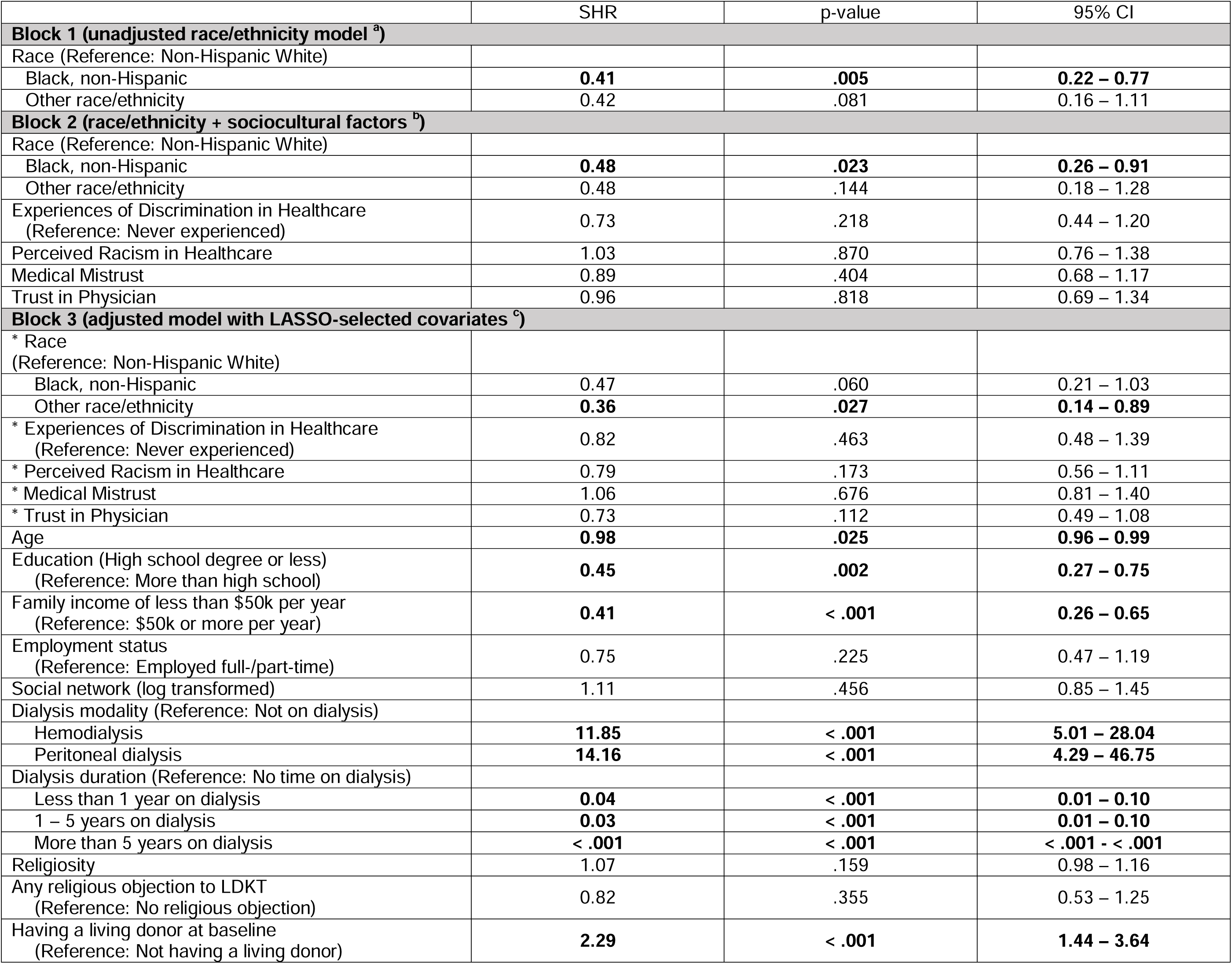
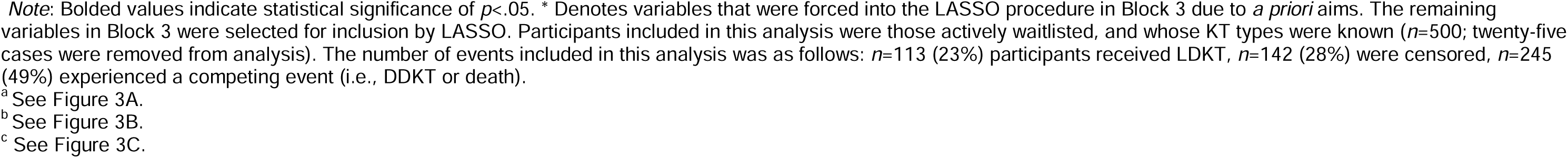
Fine-Gray proportional sub-distribution hazards models from active waitlisting to receipt of LDKT (competing events: death, DDKT)

**Table 5.** Fine-Gray proportional sub-distribution hazards models from active waitlisting to receipt of DDKT (competing events: death, LDKT)

|  | SHR | p-value | 95% CI |
| --- | --- | --- | --- |
| **Block 1 (unadjusted race/ethnicity model <sup>a</sup>)** |  |  |  |
| Race (Reference: Non-Hispanic White) |  |  |  |
| Black, non-Hispanic | **1.94** | **< .001** | **1.41 – 2.66** |
| Other race/ethnicity | **1.77** | **.038** | **1.03 – 3.05** |
| **Block 2 (race/ethnicity + sociocultural factors <sup>b</sup>)** |  |  |  |
| Race (Reference: Non-Hispanic White) |  |  |  |
| Black, non-Hispanic | **2.10** | **< .001** | **1.44 – 3.04** |
| Other race/ethnicity | 1.71 | .066 | 0.96 – 3.03 |
| Experiences of Discrimination in Healthcare<br>(Reference: Never experienced) | 1.06 | .746 | 0.75 – 1.50 |
| Perceived Racism in Healthcare | 0.83 | .097 | 0.67 – 1.03 |
| Medical Mistrust | 1.09 | .401 | 0.89 – 1.34 |
| Trust in Physician | 1.15 | .245 | 0.91 – 1.47 |
| **Block 3 (adjusted model with LASSO-selected covariates <sup>c</sup>)** |  |  |  |
| * Race (Reference: Non-Hispanic White) |  |  |  |
| Black, non-Hispanic | **1.59** | **.028** | **1.05 – 2.41** |
| Other race/ethnicity | 1.78 | .077 | 0.94 – 3.38 |
| * Experiences of Discrimination in Healthcare<br>(Reference: Never experienced) | 1.02 | .935 | 0.69 – 1.49 |
| * Perceived Racism in Healthcare | 0.88 | .269 | 0.70 – 1.10 |
| * Medical Mistrust | 1.02 | .860 | 0.81 – 1.28 |
| * Trust in Physician | 1.20 | .177 | 0.92 – 1.56 |
| Age | 1.00 | .498 | 0.98 – 1.01 |
| Education (High school degree or less)<br>(Reference: More than high school) | 1.18 | .303 | 0.86 – 1.63 |
| Family income of less than \$50k per year<br>(Reference: \$50k or more per year) | **1.51** | **.031** | **1.04 – 2.19** |
| Employment status (Unemployed)<br>(Reference: Employed full-/part-time) | 1.38 | .083 | 0.96 – 1.98 |
| Dialysis duration (Reference: No time on dialysis) |  |  |  |
| Less than 1 year on dialysis | 1.17 | .355 | 0.84 – 1.65 |
| 1 – 5 years on dialysis | **1.84** | **.004** | **1.21 – 2.81** |
| More than 5 years on dialysis | **4.50** | **.003** | **1.70 – 11.96** |
| No religious objection to LDKT<br>(Reference: Any religious objection to LDKT) | 0.97 | .854 | 0.71 – 1.32 |
| Have a living donor at baseline<br>(Reference: Does not have a living donor) | 1.00 | .997 | 0.70 – 1.42 |

Note: Bolded values indicate statistical significance of $p < .05$ . \* Denotes variables that were forced into the LASSO procedure in Block 3 due to *a priori* aims. The remaining variables in Block 3 were selected for inclusion by LASSO. Participants included in this analysis were those actively waitlisted, and whose KT types were known ( $n=500$ ; twenty-five cases were removed from analysis). The number of events included in this analysis was as follows: $n=192$ (38%) participants received DDKT, $n=142$ (28%) were censored, $n=166$ (33%) experienced a competing event (i.e., LDKT or death).
<sup>a</sup> See Figure 4A.
<sup>b</sup> See Figure 4B.
<sup>c</sup> See Figure 4C.

### Time from Active Waitlisting to DDKT

In Block 1, a greater cumulative incidence of DDKT was observed among Black participants (SHR=1.94; 95% CI: 1.41-2.66; *p*<.001) and Other race/ethnicity participants (SHR=1.77; 95% CI: 1.03-3.05; *p*=.038) compared with White participants. Black participants maintained a significantly higher cumulative incidence of DDKT receipt compared with White participants in Block 2 (SHR=2.10; 95% CI: 1.44-3.04; *p*<.001), while Other race/ethnicity participants did not (SHR=1.71; 95% CI: 0.96-3.03; *p*=.066). In Block 3, Black participants still had a significantly greater cumulative incidence of DDKT over time (SHR=1.59; 95% CI: 1.05-2.41; *p*=.028), but Other race/ethnicity participants did not (SHR=1.78; 95% CI: 0.94-3.38; *p*=.077). No statistically significant associations between sociocultural factors and DDKT receipt were identified (see Table 5 and Figure 4A-C).

**Figure 4.**
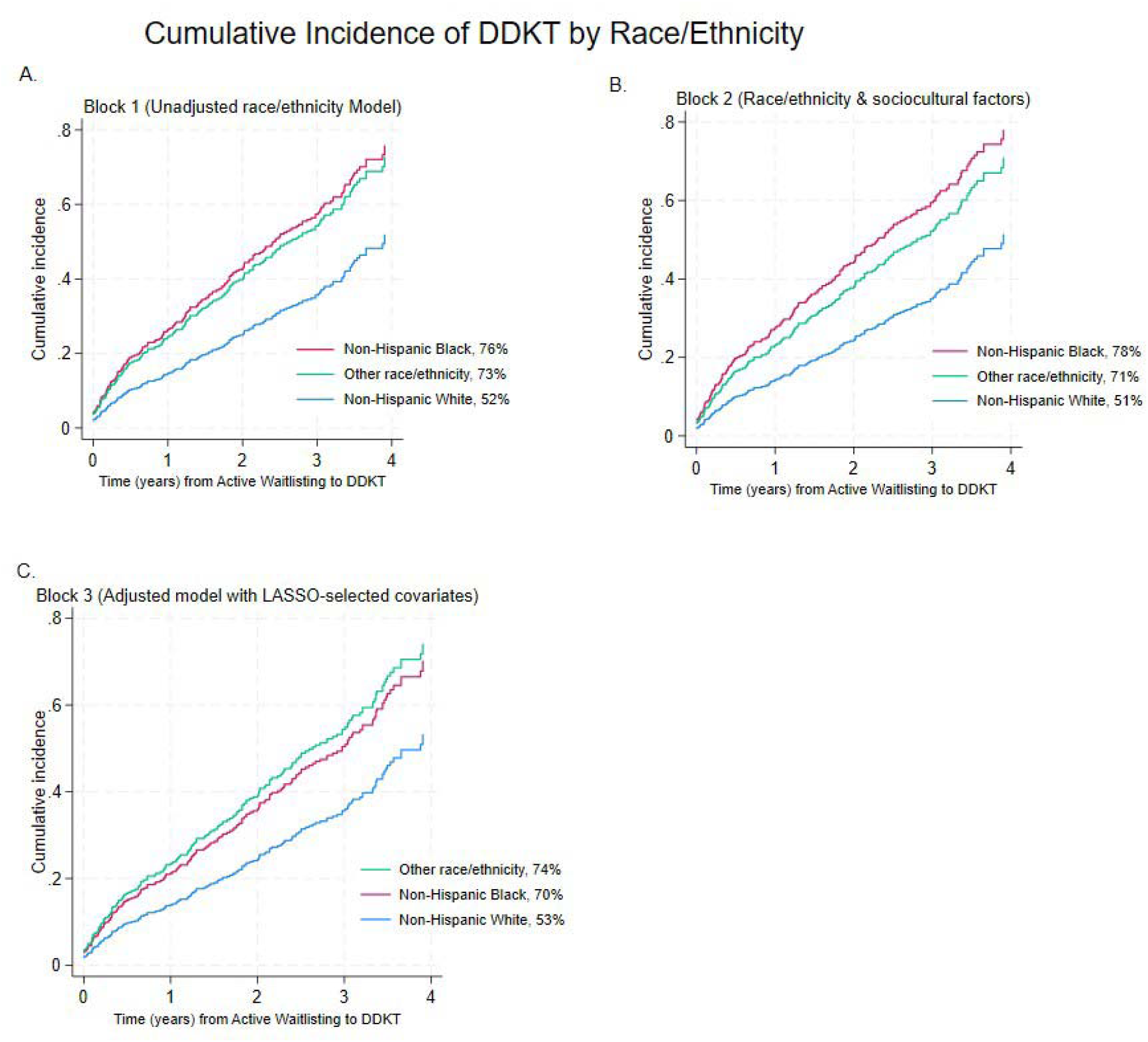
Cumulative incidence of DDKT, by race/ethnicity, estimated using Fine-Gray proportional hazard modeling (competing events: Death, LDKT): Panel A. Block 1 (Unadjusted Model); Panel B. Block 2 (Adjusted for Sociocultural Factors); Panel C. Block 3 (Fully-Adjusted Model).

## Discussion

In this exploratory secondary analysis of a prospective observational cohort of patients who underwent the KTFT approach to KT evaluation, we examined whether sociocultural factors predicted multiple KT outcomes in time-to-event models, including waitlisting, KT, and type of transplant received, and whether racial and ethnic differences existed for each KT type. We found that sociocultural factors did not predict KT receipt outcomes. Medical mistrust, however, remained significantly associated with active waitlisting even when included alongside race/ethnicity and other covariates. Additionally, we identified racial and ethnic differences in KT type (LDKT and DDKT) that persisted after accounting for sociocultural factors and other covariates, and within the context of KTFT, a systemic, clinic-level change. This work extends prior KTFT analyses by adjusting for an expanded set of covariates (e.g., sociocultural factors), and provides the first characterization of racial and ethnic variations in LDKT and DDKT receipt within this cohort.

This study is significant because it builds on the existing body of mostly cross-sectional literature^8,26,28,51^ by using a prospective cohort design to examine how sociocultural factors are associated with multiple KT outcomes in longitudinal, time-to-event models—from evaluation initiation, through KT receipt—within the context of an equity-focused intervention. Whereas prior analyses of the KTFT cohort focused solely on waitlisting and overall KT receipt and only adjusted for sociodemographic and medical factors,^19^ the present study expanded the model to include psychosocial factors, donor preference and recruitment, transplant knowledge and concerns, and sociocultural factors. In addition, we assessed these factors using a hierarchical approach to observe whether racial/ethnic associations with KT access changed after adjustment for sociocultural factors and other covariates. This systematic approach provided a more structured and comprehensive understanding of factors that may shape KT access.

Although previous studies have shown that discrimination, racism, medical mistrust, and trust in physician, are important to consider within a KT evaluation context,^7,8,18^ none have evaluated how these factors operate once major systemic barriers have been minimized. By examining the KTFT cohort, we were able to determine the influence of sociocultural factors, such as medical mistrust, which was inversely associated with active waitlisting, even when racial/ethnic disparities in waitlisting had been mitigated. Contrary to our expectations, we found that sociocultural factors were not significantly associated with any KT outcome, suggesting either that their influence may occur earlier in the chronic kidney disease (CKD) trajectory, or that KTFT may have mitigated their influence beyond waitlisting.

Furthermore, although we again observed no racial differences in overall KT receipt, which is consistent with KTFT’s goal and our prior study,^19^ we found that racial and ethnic differences persisted to some degree in the KT type received. White participants were more likely to receive LDKT, but Black and Other race/ethnicity participants more commonly received DDKT. Interestingly, racial/ethnic differences in KT type shifted with model adjustment. For example, Black-White differences in LDKT receipt attenuated after accounting for additional factors, whereas significant differences emerged between White and Other race/ethnicity participants only after adjustment. Notably, Black participants remained significantly more likely to receive DDKT compared with White participants even in the fully-adjusted model, whereas the difference between Other race/ethnicity and White participants was reduced beyond the univariate model. These findings clarify that interventions addressing disparities in KT receipt must consider LDKT and DDKT separately, because there may be masked differential effects in KT type. Further, results suggest that, rather than sociocultural factors, it may be that broader contextual factors, such as access to living donors, influence KT outcomes, even within an intervention designed to promote KT access.

Patients in this KTFT cohort benefitted from a streamlined approach to KT evaluation.

Still, the persistent association between medical mistrust and waitlisting, as well as the sustained racial differences in type of KT received, underscore the limitations of relying on clinic-level, equity-focused interventions alone. In one prior analysis of the KTFT cohort, we found that patients reported improvements in perceived discrimination, racism, and medical mistrust after completing KTFT, but Black participants’ trust in physicians declined.^18^ Taken together with our current findings, this points to the need for systemic interventions to be paired with more personalized strategies that intentionally build trust and address patients’ prior experiences at both interpersonal *and* institutional levels. Although KTFT addressed a critical structural barrier (i.e., lengthy KT evaluation process), additional strategies at the individual-and interpersonal-levels, like culturally-concordant peer support to improve patient understanding of optimal KT treatments, or provider trainings to enhance shared decision-making processes, may build trust in healthcare providers and systems, and support equitable outcomes.^52^ Notably, in this same cohort, a culturally-tailored educational intervention did not improve KT-related outcomes overall,^53^ underscoring that education alone may be insufficient when patients face broader systemic barriers, or have limited agency to act on new knowledge. Embedding trust-building and patient-empowerment strategies within systemic interventions, like KTFT, may close remaining gaps in KT access and equity. Moreover, interventions introduced even earlier in the CKD continuum may also meaningfully contribute to reduced disparities. For example, timely referral to nephrology, early nephrology engagement, and education about kidney failure treatments and KT options may address inequities that manifest long before KT evaluation begins.^54^

## Limitations

In the present study, we conducted a secondary analysis of a prospective observational cohort. Because we did not conduct a randomized-controlled trial, or include a comparison group that was not exposed to the KTFT program, we cannot draw any causal inferences regarding the observed associations among the key outcomes. Baseline measures were collected once, prior to the initial KT evaluation appointment. Past research suggests sociocultural factors may evolve over time,^18^ so it is possible we were limited in our ability to detect associations with longer-term outcomes like KT receipt. Additionally, baseline measures were either self-administered or obtained with the help of an interviewer. As such, responses were subject to social acceptability bias, either due to the presence of the interviewer, or for fear their responses would influence their candidacy for active waitlisting—despite the fact participants were assured their responses would not affect their healthcare. Also, this study was conducted at a single KT clinic in the United States, thus limiting generalizability of findings. The Other race/ethnicity group was small (7% of sample) and heterogeneous, so findings regarding this group should be interpreted with caution. Nevertheless, the present study had a relatively diverse sample of patients from urban and rural backgrounds, and we were able to examine multiple KT outcomes prospectively, and over long-term follow-up.

## Conclusions

In this prospective observational cohort study of KT candidates who underwent a streamlined approach to KT evaluation, we observed that medical mistrust remained a significant barrier to active waitlisting. We also identified racial/ethnic differences in LDKT and DDKT receipt. Our analytic approach accounted for a broad range of relevant factors, including psychosocial, donor recruitment and preferences, and transplant knowledge and concerns, offering a comprehensive understanding of what may influence KT access. These findings suggest that although clinic-level interventions like KTFT are critical to address structural barriers, additional measures can reduce racial and ethnic disparities in KT outcomes.

Personalized strategies that build trust and address patients’ prior experiences in healthcare at the individual-and interpersonal-levels may be necessary alongside systemic efforts. Future research should explore the impact of multi-level and upstream interventions on KT outcomes.

## Data Availability

Dr. Larissa Myaskovsky had full access to all the data in the study and takes responsibility for the integrity of the data and the accuracy of the data analysis. The data are not publicly available due to privacy or ethical restrictions.

## Acknowledgment Section

Disclosure of potential conflicts of interest

None to disclose.

## Author Contributions

Dr. Vélez-Bermúdez is the first author. Dr. Myaskovsky is the senior author.

*Concept and design:* Vélez-Bermúdez, Dew, and Myaskovsky.

*Acquisition, analysis, or interpretation of data*: Vélez-Bermúdez, Leyva, Zhu, Dew, and Myaskovsky.

*Drafting of the manuscript:* Vélez-Bermúdez and Myaskovsky.

*Critical review of the manuscript for important intellectual content:* All authors.

*Statistical analysis*: Vélez-Bermúdez, Leyva, Zhu.

*Obtained funding*: Myaskovsky.

*Supervision:* Myaskovsky, Zhu, Unruh, and Dew.

## Acknowledgments

The authors thank the individuals who participated in this study for their contributions to this research.

## Abbreviations

KT: Kidney transplantation
KTFT: Kidney Transplant Fast Track
KAS: Kidney Allocation System
LDKT: Living donor kidney transplantation
DDKT: Deceased donor kidney transplantation
CKD: Chronic kidney disease

