## Supplemental Material for "Healthcare-related sociocultural factors, racial disparities, and kidney transplant outcomes in the Kidney Transplant Fast Track program"

**Supplemental Contents**

**Supplemental Table 1.** Strengthening the Reporting of Observational Studies in Epidemiology Checklist

**Supplemental Table 2.** Baseline characteristics and potential predictors

**Supplemental Material References.**

**Supplemental Table 1.** Strengthening the Reporting of Observational Studies in Epidemiology Checklist^1^

|  | Item No | Recommendation | Page No |
| --- | --- | --- | --- |
| **Title and abstract** | 1 | (*a*) Indicate the study’s design with a commonly used term in the title or the abstract | 2 |
|  |  | (*b*) Provide in the abstract an informative and balanced summary of what was done and what was found | 2 |
| Introduction | | | |
| Background/rationale | 2 | Explain the scientific background and rationale for the investigation being reported | 3-5 |
| Objectives | 3 | State specific objectives, including any prespecified hypotheses | 5 |
| Methods | | | |
| Study design | 4 | Present key elements of study design early in the paper | 5-6 |
| Setting | 5 | Describe the setting, locations, and relevant dates, including periods of recruitment, exposure, follow-up, and data collection | 5-6 |
| Participants | 6 | (*a*) Give the eligibility criteria, and the sources and methods of selection of participants. Describe methods of follow-up | 5-6 |
|  |  | (*b*) For matched studies, give matching criteria and number of exposed and unexposed | N/A |
| Variables | 7 | Clearly define all outcomes, exposures, predictors, potential confounders, and effect modifiers. Give diagnostic criteria, if applicable | 6-7 |
| Data sources/ measurement | 8* | For each variable of interest, give sources of data and details of methods of assessment (measurement). Describe comparability of assessment methods if there is more than one group | 6-7, Supplemental Table 2 |
| Bias | 9 | Describe any efforts to address potential sources of bias | 7-9 |
| Study size | 10 | Explain how the study size was arrived at | 6 |
| Quantitative variables | 11 | Explain how quantitative variables were handled in the analyses. If applicable, describe which groupings were chosen and why | 6-9 Supplemental Table 2 |
| Statistical methods | 12 | (*a*) Describe all statistical methods, including those used to control for confounding | 7-9 |
|  |  | (*b*) Describe any methods used to examine subgroups and interactions | 7-9 |
|  |  | (*c*) Explain how missing data were addressed | 7-9, Table 1-5 |
|  |  | (*d*) If applicable, explain how loss to follow-up was addressed | 5-9, Table 1-5 |
|  |  | (*e*) Describe any sensitivity analyses | N/A |
| Results | | |  |
| Participants | 13* | (a) Report numbers of individuals at each stage of study—e.g., numbers potentially eligible, examined for eligibility, confirmed eligible, included in the study, completing follow-up, and analyzed | 6, Tables 2-5 |
|  |  | (b) Give reasons for non-participation at each stage | Tables 2-5, |
|  |  | (c) Consider use of a flow diagram | 6 |
| Descriptive data | 14* | (a) Give characteristics of study participants (e.g., demographic, clinical, social) and information on exposures and potential confounders | 9, Table 1 |
|  |  | (b) Indicate number of participants with missing data for each variable of interest | Table 1 |
|  |  | (c) Summarize follow-up time (e.g., average and total amount) | Figure 1-4 |
| Outcome data | 15* | Report numbers of outcome events or summary measures over time | Tables 2-5 |

| **Supplemental Table 2. Baseline characteristics and potential predictors collected before the initial KT evaluation appointment** | | | |
| --- | --- | --- | --- |
| **Variables** | | **Description** | **Coding/ Range/ Cronbach’s α (if applicable)** |
| **Demographic and sociocultural factors at baseline included in all multivariable analyses** | | | |
|  | Race/ethnicity | Grouped into the following categories:  1. Non-Hispanic White  2. Non-Hispanic Black  3. Other (includes American Indian or Alaska Native, Asian, Hispanic or Latine, and/or Native American or Other Pacific Islander) | Categorical, 3 categories |
|  | Experience of discrimination in medical settings^2,3^ | 7 items assessing perceived discrimination in a healthcare setting; original range: 1 (never) to 5 (always) | Dichotomized for analysis: “ever experienced discrimination” versus “never experienced discrimination”  Cronbach’s α = 0.88 |
|  | Perceived racism^4,5^ | 4 items assessing patients’ belief that racism is common in healthcare | Mean score calculated for analysis  Continuous  Responses and potential range: 1 (strongly disagree) to 5 (strongly agree)  Cronbach’s α = 0.75 |
|  | Medical mistrust^5,6^ | 7 items assessing beliefs that patient’s hospital is trustworthy, competent, and acting in their best interests | Mean score calculated for analysis  Continuous  Responses and potential range: 1 (strongly disagree) to 5 (strongly agree);  Cronbach’s α = 0.78 |
|  | Trust in physicians^7^ | 11 items assessing patients’ trust in their physician | Mean score calculated for analysis  Continuous  Responses and potential range: 5 (totally disagree) to 1 (totally agree)  Cronbach’s α = 0.85 |
| **Demographic characteristics, tested in LASSO for inclusion in subsequent multivariable analyses** | | | |
|  | Age | Age at baseline interview | Continuous  Range in present sample: 20 – 88 |
|  | Sex | Men, women | Dichotomous |
|  | Education | Patient-reported using the following choices:  1. Less than high school  2. Some high school  3. High school graduate  4. Some college  5. College degree  6. Graduate degree | Dichotomized for analysis: “Less than or equal to high school” versus “Greater than high school education” |
|  | Household Income | Patient-reported using the following choices:  1. Under $15,000 2. $15,000 - $24,999 3. $25,000 - $49,999 4. $50,000 - $74,999 5. $75,000 - $100,000 6. Over $100,000 | Dichotomized for analysis: “Less than $50,000” versus “Greater than or equal to $50,000” |
|  | Marital status | Patient-reported using the following categories:  1. Single (never married)  2. Separated or Divorced  3. Widowed  4. Married  5. Domestic Partnership | Dichotomized for analysis: “Married/In a Domestic partnership” versus “Single/Separated or Divorced/Widowed” |
|  | Insurance status | Patient-reported and grouped into the following categories:  1. Private insurance  2. Private/public mix  3. Public insurance only | Categorical, 3 categories |
|  | Employment status | Patient-reported using the following categories:  1. Yes, full time  2. Yes, part time  3. No, unemployed | Dichotomized for analysis: “Full- or part-time employment” versus “unemployed” |
|  | Number in social network (i.e., “network of potential living donors”) | Patient-reported number of the network of potential living donors available for evaluation was determined by asking participants to indicate how many living relatives and friends they had aged 18–70 years of age. | Continuous  Range in present sample: 0 – 150 |
| **Medical factors, tested in LASSO for inclusion in subsequent multivariable analyses** | | | |
|  | Body mass index (BMI) | Calculated from medical record, value was squared in analysis to adjust for skewness | Continuous  Range in present sample: 13.49 – 53.84 |
|  | Charlson Comorbidity Index^8^ | From medical record data: weighted score reflecting the number and severity of co-morbid health conditions | Continuous: 0 (no comorbidities) to 33 (a higher number of comorbidities or more serious comorbidities)  Range in present sample: 2 – 11 |
|  | Dialysis duration | From medical record:  1. 0 years on dialysis  2. <1 year on dialysis  3. 1-<5 years on dialysis  4. > 5 years | Categorical, 4 categories |
|  | Dialysis type | From medical record: 1. Hemodialysis  2. Peritoneal dialysis  3. No dialysis | Categorical, 3 categories |
|  | Burden of Kidney Disease^9^ | Subscale that measures the extent to which kidney disease interferes with the respondent’s life. | Mean score calculated for analysis  Continuous  Responses and potential range: 1 (Definitely false) to 5 (Definitely true)  Cronbach’s α = 0.76 |

| **Psychosocial characteristics, tested in LASSO for inclusion in subsequent multivariable analyses** | | | | | |
| --- | --- | --- | --- | --- | --- |
|  | Social support^10,11^ | | 12-item Interpersonal Support Evaluation List (ISEL-12) - assessed perceived availability of 3 separate functions of social support: "tangible," "appraisal,” and "belonging". | | Continuous  Range in present sample: 12 – 48  Cronbach’s α = 0.86 |
|  | Anxiety ^12^ | | 6-item Brief Symptom Inventory (BSI) Anxiety subscale where participants report the extent to which they indicate how bothered or distressed they have felt in the past 2 weeks by several symptoms. Responses ranged from 1 (“Not at all”) to 5 (“Extremely”). Responses were dichotomized to reflect “No anxiety” or “Moderate to severe anxiety.” | | Dichotomized for analysis: “No anxiety” versus “Moderate to severe anxiety”  Cronbach’s α = 0.83 |
|  | Depression ^12^ | | 6-item Brief Symptom Inventory (BSI) Depression subscale where participants report the extent to which they indicate how bothered or distressed they have felt in the past 2 weeks by several symptoms. Responses ranged from 1 (“Not at all”) to 5 (“Extremely”). Responses were dichotomized to reflect “No depression” or “Moderate to severe depression.” | | Dichotomized for analysis: “No depression” versus “Moderate to severe depression”  Cronbach’s α = 0.83 |
|  | Family loyalty^13^ | | 16 items assessing participants’ loyalty and mutual support regarding the family | | Mean score calculated for analysis  Continuous  Responses and potential range: 1 (totally disagree) to 5 (totally agree);  Cronbach’s α = 0.84 |
|  | Overall Religiosity^14^ | | 2 items assessing the importance and influence of religious beliefs in a participants’ life | | Mean score calculated for analysis  Continuous  Responses and potential range: 1 (not at all important) to 9 (very important);  Cronbach’s α = 0.82 |
|  | Religious objections to LDKT^15^ | | Revised subscale of the 8 item Organ Donation Attitude Survey (ODAS). From these questions, we categorized respondents into 3 groups:  1. No objection – “disagree” or “strongly disagree” with all religious objections to transplant  2. Neutral – combination of “disagree,” “strongly disagree” and “not sure” toward religious objection to transplant  3. Any objection – “agree” or “strongly agree” with any religious objection to transplant | | Dichotomized for analysis: “any religious objection to LDKT” versus “no religious objection to LDKT”  Cronbach’s α = 0.72 |
|  | Health Literacy^16^ | | A three-item measure assessing participants’ understanding about their own health information, with scores ranging from 1 (“Always,” “Extremely”) to 5 (“Never,” “Not at all”). Higher scores reflect higher literacy. The measure was scored by taken the mean score of each item. In this sample, health literacy scores ranged from 1 – 5. | | Mean score calculated for analysis  Continuous  Cronbach’s α = 0.67 |
| **Transplant knowledge and concerns, tested in LASSO for inclusion in subsequent multivariable analyses** | | | | | |
|  | Transplant knowledge^17^ | Participants were assessed on their knowledge about transplant using a 19-item KT Knowledge Survey. Items 1 – 8 were multiple choice, and items 9 – 19 were “True” or “False.” Each item answered correctly adds 1 to total score. | | Continuous  Range in present sample: 0 – 18  Cronbach’s α = 0.84 | |
|  | Number of learning activities^18^ | Participants reported the type and number of KT-related learning activities (e.g., reading brochures, online research) with greater numbers indicating engagement in more learning activities. | | Continuous  Range in present sample: 0 – 4  Cronbach’s α = 0.54 | |
|  | Hours engaged in learning activities^18^ | Participants reported the amount of time spent in KT-related learning activities (e.g., reading brochures, online research). | | Categorized for present analysis:  1: 0 – 2 hours of learning  2: Greater than 2 hours, less than or equal to 5 hours  3: Greater than 5 hours of learning  Cronbach’s α = 0.59 | |
|  | Transplant concerns | In a 24-item assessment, participants reported which of 24 common transplant-related concerns were most important in influencing their decision to pursue transplant. | | Sum score; continuous.  Item responses ranged from 1 (“Not important”) to 5 (“Extremely important”) Range in present sample: 16 – 60  Cronbach’s α = 0.80 | |
| **Donor recruitment and preference variables, tested in LASSO for inclusion in subsequent multivariable analyses** | | | | | |
|  | Donation preference | Patient-reported using the following categories:  1. Transplant from a deceased donor  2. Transplant from a living donor | | Dichotomous | |
|  | Having a living donor at baseline | Patient-reported using the following categories:  0 = No  1 = Yes | | Dichotomous | |
|  | Willingness to accept a living donor | Patient-reported using the following categories:  0 = No  1 = Yes | | Dichotomous | |
|  | Willingness to ask for a living donor | Patient-reported using the following categories:  0 = No  1 = Yes | | Dichotomous | |

**Supplemental Material References**

11.	Cohen S, Zdaniuk B. ISEL 12 Psychometric Properties.

12.	Derogatis LR. Brief symptom inventory. European Journal of Psychological Assessment. Published online 1975.
